# Informatic tools for diagnosis in dentistry. A compilation review

**DOI:** 10.1101/2024.05.05.24306900

**Authors:** Alain Manuel Chaple Gil, Solanch de la Caridad Damas Heredia, Kelvin I. Afrashtehfar

## Abstract

The objective of this study was to compile the computer tools available in the scientific literature aimed at diagnosis in dentistry. A scoping review was conducted using *PubMed*, *Scopus*, and *Web of Science*. Were include, original researches type articles, Articles that reported the usefulness of a computer/technological tool that helps diagnosis in dental practice, Articles published in the last 20 years (period 2004-2024) and written in English and Spanish. Online tool *Rayyan®* was used to establish homogeneity in the review of the authors on a single online platform where they had access and could centralize the results. Variables were extracted from the articles included in the study. In total, 12648 records were retrieved from the database. After decantation, 39 reports described 36 computer tools used for diagnosis in dentistry. More informatic tools related to "Restorative Dentistry’ have been developed than the rest of the specialties 14 (40%). Python was the predominant programming language, 83.3% of the tools were validated, and 27.8% were free. Informatics tools in dentistry enhance the diagnosis and treatment planning. However, a robust regulatory framework is required for validation prior to clinical implementation. Continuous training of dental professionals using these technologies is crucial to maximize their benefits and ensure optimal patient care. More research is needed to explore the potential of informatics applications in dentistry, their integration into existing health systems, and their accessibility in resource-limited areas.

## INTRODUCTION

Dentistry, like other medical disciplines, has undergone a significant digital transformation over the past few decades. The incorporation of digital technologies has improved the efficiency, precision, and scope of dental procedures, positively affecting both professionals and patients. However, the availability of a wide range of digital tools for diagnosis in dentistry poses a significant challenge due to the mercantilist stocks of the most widely used.^(1, 2)^

A health informatics tool is a system or platform designed to acquire, process, interpret, and use healthcare data efficiently.^(3)^ These tools aim to improve patient safety by identifying the hazards and risks associated with health information technology.^(4)^ They play a crucial role in improving health service delivery by ensuring that the information reaches the right person at the right time. Such tools help in decision making, access to information, disease prevention, and improved communication, allowing patients and their families to manage their health effectively. These tools span various technologies, such as wearable devices, passive monitoring tools, and smart homes, providing personalized interventions based on patient-generated health data.^(5)^

Computer tools are increasingly used in dental practice for diagnostic purposes. These tools use intelligent systems and machine learning algorithms to analyze dental images and detect various dental conditions such as periodontal disease, alveolar bone loss, and dental pathologies. The use of deep convolutional neural networks (CNN) has yielded promising results in accurately detecting and classifying these conditions based on radiographic images.^(6)^ In addition, machine vision algorithms applied to high-resolution images can help diagnose periapical lesions and plan endodontic treatment.^(7)^ Computerized dental office systems also contribute to improved efficiency and diagnostic tasks, such as early detection of cavities, through image processing software and digital X-rays.^(8)^ In addition, computer systems that implement neural networks can process dental images to detect and diagnose pathological conditions, providing a graphical user interface for visualization and correlation with dental medical records.^(9)^ These informatics tools offer valuable support for dental diagnosis, treatment planning, and patient education, and improve the overall quality of dental care.^(6, 8, 10)^

The proliferation of computer tools for diagnosis in dentistry presents opportunities, but also generates a series of questions and difficulties for dentists.^(11, 12)^ There is a wide variety of these, from radiographic image analysis software to treatment design and planning systems, to augmented reality applications and intraoral scanners. This diversity makes it difficult to identify the most appropriate tools for each case.^(12)^

The identification and selection of appropriate informatics tools for diagnosis in dentistry is crucial to improve diagnostic accuracy, as they can provide additional and more accurate information for diagnosis, allowing for better treatment planning.^(13)^ They optimize workflow by being able to streamline diagnostic processes, improving efficiency and productivity and can help visualize, explain problems and treatment options to the patient, improving understanding and decision-making.^(14)^

### Purpose Definition

The purpose of this review was to identify the informatics tools reported in the scientific literature that have been tested to aid diagnosis in dental practice. Based on this approach, the research problem is shown through the following question: What are the computer tools reported by science that could help establish diagnoses in dental practice?

## OBJECTIVE

The objective of this study was to compile the informatics tools available in the scientific literature aimed at diagnosis in dentistry.

## METHODOLOGY

A scoping review was conducted between January and April 2024. The *PubMed*, *Scopus*, and *Web of Science* (WOS) databases were used for this development. The reporting guidelines for literature reviews described in the PRISMA guidelines were followed.

### Inclusion criteria

Were include, original researches type articles, Articles that reported the usefulness of a computer/technological tool that helps diagnosis in dental practice, Articles published in the last 20 years (period 2004-2024) and written in English and Spanish.

### Exclusion Criteria

Review articles of any kind (narrative, systematic, and meta-analyses), letters, case presentations, books, theses, *preprints*, articles describing computer tools for diagnosis unrelated to stomatology, articles describing generic software for the visualization of images taken with CAD-CAM or similar equipment whose validations are frankly demonstrated, articles written in languages other than English and Spanish and Articles that did not specify a name for the tool they report were excluded.

### Search strategy

A search was carried out using advanced formulations on the three platforms described in Table 1. Entered keywords combined with wildcards and Boolean operators were displayed.

**Table 1.** Formulations of the search by database used.

| Database | Formulation |
| --- | --- |
| Pubmed | ((application[Title/Abstract]) OR (program[Title/Abstract]) OR (tool[Title/Abstract]) OR (informatic[Title/Abstract]) OR (computer[Title/Abstract]) OR (digital[Title/Abstract]) OR (apk[Title/Abstract]) OR (system[Title/Abstract]) OR (automatic[Title/Abstract])) AND ((dental[Title/Abstract]) OR (dentistry[Title/Abstract])) AND ((diagnosis[Title/Abstract]) OR (diagnoses[Title/Abstract])) |
| Scopus | (TITLE-ABS-KEY (program*) OR TITLE-ABS-KEY (application*) OR TITLE-ABS-KEY (tool*) OR TITLE-ABS-KEY (comput*) TITLE-ABS-KEY (informatic*) OR TITLE-ABS-KEY (apk) OR TITLE-ABS-KEY (system*) OR TITLE-ABS-KEY (digital*) OR TITLE-ABS-KEY (automa*)) AND (TITLE-ABS-KEY (dent*)) AND (TITLE-ABS-KEY (diagnos*)) |
| WOS | (TI=(program*) OR TI=(application*) OR TI=(tool*) OR TI=(comput*) OR TI=(informatic*) OR TI=(apk) OR TI=(system*) OR TI=(digital*) OR TI=(automa*)) AND (TI=(dent*)) AND (TI=(diagn*)) |

### Methodology for the selection of articles

In the databases where the searches were performed, automated filters were applied to discard articles that were not of the type described in the inclusion criteria and to establish the publication time range.

Records extracted from the databases were exported to *Clartivates* Analytics EndNote 21. In this same system, duplicate articles that did not meet the eligibility criteria were decanted at first glance.

Calibration was performed among the authors to evaluate the articles to be selected. The degree of coincidence of the evaluations made by the reviewers was determined using Orwin’s method of 1994, and a Kappa statistic was performed to measure the agreement among the reviewers who would make simple decisions about inclusion/exclusion. Kappa values between 0.40 and 0.59 were considered to reflect acceptable agreement, 0.60 to 0.74 to be an adequate agreement, and 0.75 or more to reflect excellent agreement.

The online tool *Rayyan®* was used to establish homogeneity in the authors’ review on a single online platform where they had access and could centralize the results. When the two reviewers did not agree to the decision to include or exclude a registry, a third party intervened to make the final decision.

First, duplicate reports written in languages other than English and Spanish were discarded, applying obvious inclusion/exclusion criteria such as type of document and year of publication. Subsequently, articles that did not meet the criteria were discarded based on their titles and abstracts. Reports with titles and abstracts that were doubtful of being discarded or were not discarded were read to determine this. Once those articles that did not meet the criteria were discarded, the included articles were read in detail to extract the necessary data related to the research variables.

### Variables

The following variables were extracted from each article included in the study: name and surname of the first author, year of publication, Article Title, Journal in which it was published, citation details (including volume, number, and pages), the URL or DOI of the document (preferably the DOI), name of the tool described, country where it was developed, Tool Platform (this refers to the programming language in which the tool was developed), environment (Windows, Linux, Unix, Android, iOS, or other), dental specialty to which it pays, a brief description of the usefulness of the tool, tool validation (dichotomous yes or no), open source (dichotomous yes or no), and usage charge (dichotomous yes or no). For the Payment model, *full payment* is considered: users pay a one-time price to access the app and all its functionalities, a recurring fee (monthly, yearly, etc.) to access the application and its functionalities, or a license is purchased to use the application for a certain period or in perpetuity. *Freemium*: The app can be used for free, but it offers optional in-app purchases to unlock additional features, exclusive content, or to remove advertising. In addition, it was included that they were free versions with basic functionalities and paid versions with more advanced functionalities. *Trial model*: This offers a free trial period during which all features can be accessed. After the trial period, the user must pay to continue using the app and *Free Model*: Completely free, without any payment or restrictions.

In addition, the URL through which the user manual and/or instructions for the installation and use of each tool can be downloaded was included, if available.

### Data management

The records of the variable data extracted from the included articles were captured in a Microsoft® Excel® spreadsheet to later favor the analysis and preparation of tables and figures for better understanding.

In cases in which the articles were not sufficiently explicit, the authors were contacted via email for more information. Respondents increased the scope of the information for this research. Articles by authors who did not respond within a reasonable period of time and the information in the articles that did not appear sufficiently for data collection were excluded.

The database of the articles included in the study with all the details collected according to the variables is located in the Mendeley Data repository (https://doi.org/10.17632/yx9jts8mv7.1), which advocates for the reproducibility, sharing, and systematicity of data according to the principles of Open Science.

### Ethical aspects

The protocol for this study was approved by the Scientific Council of the "Ana Betancourt" Dental Clinic in Havana, Cuba.

## RESULTS

The calibration of the authors who reviewed the articles included in the study was excellent in all cases. The search was conducted on January 28 2024. In the primary search, 3397 records were established in Pubmed, 7563 in *Scopus*, and 221 in WOS, for a total of 12648. After applying the filter of publication time (2004–2024) and the type of article, there were 46 results in *PubMed* (only for *clinical trials* and *randomized controlled trial articles*), 5678 in *Scopus*, and 162 in WOS, resulting in 5886 records.

After removing duplicates, 4159 articles remained, and applying the inclusion criteria of article type 3937 and excluding articles in languages other than English and Spanish, 3655 entries remained. (Figure 1)

**Figure 1.**
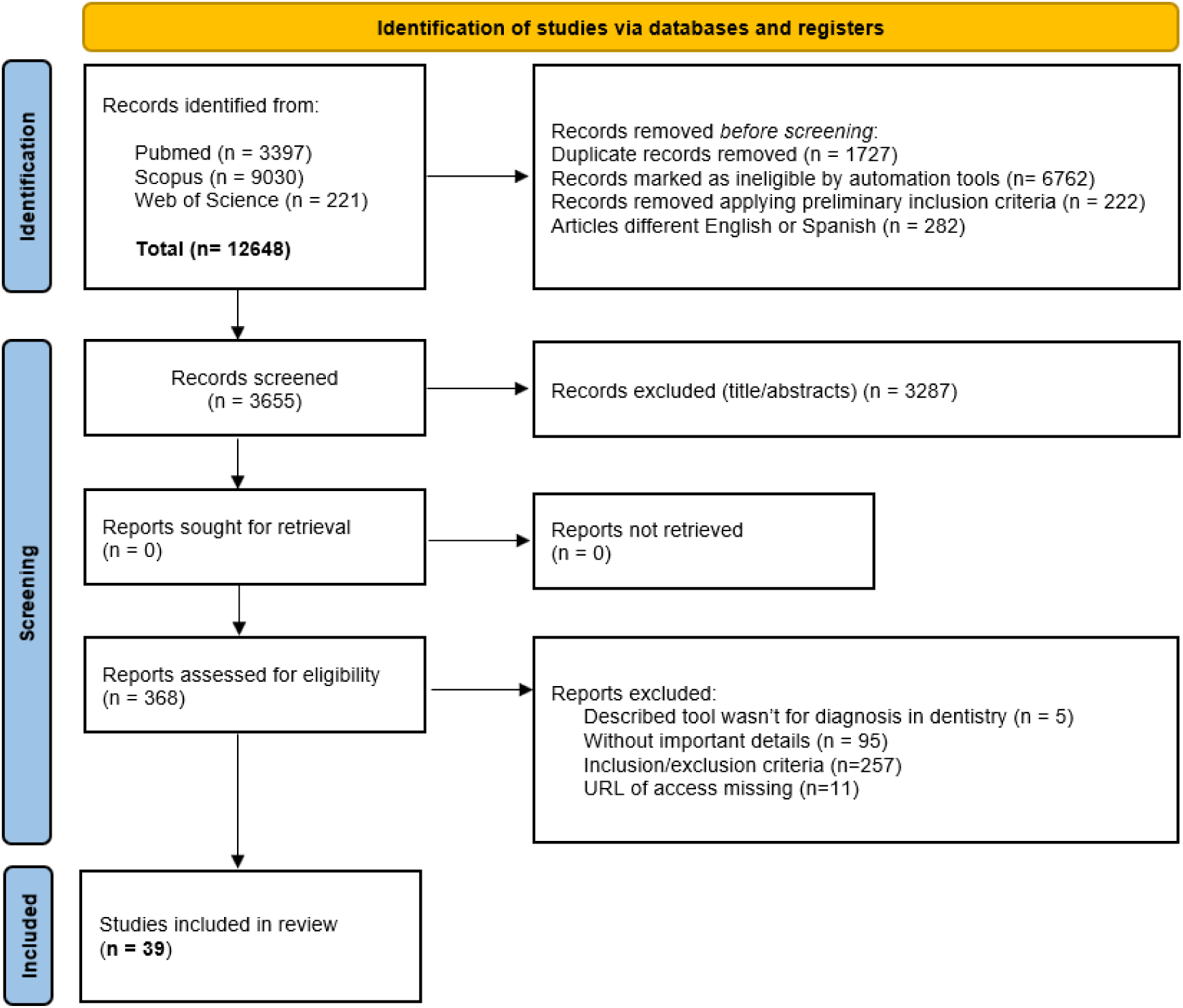
PRISMA flow diagram for systematic reviews. (**Source**: Developed by authors)

When examining the titles and abstracts of the remaining articles, 3287 articles were excluded, leaving 368 reports that were downloaded in full text and excluded five for not exactly describing a telematic tool for diagnosis in dentistry, 95 were excluded because they lacked the necessary details for the adequate collection of information, another 11 were discarded for loss of access through URLs, and 257 were not strictly complying with the criteria inclusion/exclusion. At the end of the process, 39 reports described 36 computer tools used for diagnosis in dentistry. (Figure 1)

Table 2 shows the list of articles included in the study with a predominance of articles published in the International Journal of Environmental Research and Public Health”^(15, 16, 17, 18)^ with 4 reports for 10.3% of the total, followed by the "Diagnostics”^(19, 20, 21)^ with three (7.7%) reports. Only O’Toole^(22, 23)^ matched two (5.1%) articles as the first author. The years of publication of the articles ranged from 2015 to 2023, with 2022 predominating^(17, 18, 21, 24, 25, 26, 27, 28, 29, 30)^ and 10 (25.6%) reports.

**Table 2.** List of articles included in the study with the first author’s surname, year of publication, title, and journal in which they were published.

| # | 1st Author | Year | Article Title | Journal |
| --- | --- | --- | --- | --- |
| 1 | Abd El Galil<sup>(31)</sup> | 2021 | Evaluation of two computer-aided design software on the adaptation of digitally constructed maxillary complete denture | Journal of Indian Prosthodontic Society |
| 2 | Abuabara<sup>(32)</sup> | 2023 | Evaluation of Endo 10 mobile application as diagnostic tool in endodontics | J Clin Exp Dent |
| 3 | Alalharith<sup>(15)</sup> | 2020 | A deep learning-based approach for the detection of early signs of gingivitis in orthodontic patients using faster region-based convolutional neural networks | International Journal of Environmental Research and Public Health |
| 4 | Altukroni<sup>(33)</sup> | 2023 | Detection of the pathological exposure of pulp using an artificial intelligence tool: a multicentric study over periapical radiographs | BMC Oral Health |
| 5 | Amorim<sup>(24)</sup> | 2022 | Smartphone and computer cephalometric analysis: A trueness and precision study | Revista Portuguesa de Estomatologia, Medicina Dentaria e Cirurgia Maxilofacial |
| 6 | Anacleto<sup>(34)</sup> | 2019 | Superimposition of 3d maxillary digital models using open-source software | Dental Press Journal of Orthodontics |
| 7 | Barreto<sup>(35)</sup> | 2016 | Reliability of digital orthodontic setups | Angle Orthodontist |
| 8 | Baydar<sup>(19)</sup> | 2023 | The U-Net Approaches to Evaluation of Dental Bite-Wing Radiographs: An Artificial Intelligence Stud | Diagnostics |
| 9 | Bonfanti-Gris<sup>(25)</sup> | 2022 | Evaluation of an Artificial Intelligence web-based software to detect and classify dental structures and treatments in panoramic radiographs | Journal of Dentistry |
| 10 | Cavalcante<sup>(16)</sup> | 2020 | Computing and oral health: Mobile solution for collecting, data analysis, managing and reproducing epidemiological research in population groups | International Journal of Environmental Research and Public Health |
| 11 | Cui<sup>(36)</sup> | 2021 | TSegNet: An efficient and accurate tooth segmentation network on 3D dental model | Medical Image Analysis |
| 12 | Dayi<sup>(20)</sup> | 2023 | A Novel Deep Learning-Based Approach for Segmentation of Different Type Caries Lesions on Panoramic Radiographs | Diagnostics |
| 13 | De Angelis<sup>(17)</sup> | 2022 | Artificial Intelligence: A New Diagnostic Software in Dentistry: A Preliminary Performance Diagnostic Study | International Journal of Environmental Research and Public Health |
| 14 | Di Fede<sup>(37)</sup> | 2023 | Doctoral: A smartphone-based decision support tool for the early detection of oral potentially malignant disorders | Digital Health |
| 15 | do Vale Voigt<sup>(38)</sup> | 2020 | DSDapp use for multidisciplinary esthetic planning | Journal of Esthetic and Restorative Dentistry |
| 16 | Estai<sup>(39)</sup> | 2017 | End-user acceptance of a cloud-based teledentistry system and Android phone app for remote screening for oral diseases | Journal of Telemedicine and Telecare |
| 17 | Fatima<sup>(40)</sup> | 2023 | Deep Learning-Based Multiclass Instance Segmentation for Dental Lesion Detection | Healthcare (Switzerland) |
| 18 | Fazio<sup>(18)</sup> | 2022 | LinguAPP: An m-Health Application for Teledentistry Diagnostics | International Journal of Environmental Research and Public Health |
| 19 | Gullberg<sup>(26)</sup> | 2022 | The challenge of applying digital image processing software on intraoral radiographs for osteoporosis risk assessment | Dentomaxillofacial Radiology |
| 20 | Haron<sup>(41)</sup> | 2020 | M-Health for Early Detection of Oral Cancer in Low- and Middle-Income Countries | Telemedicine and e-Health |
| 21 | Jiang<sup>(42)</sup> | 2021 | RDFNet: A Fast Caries Detection Method Incorporating Transformer Mechanism | Computational and Mathematical Methods in Medicine |
| 22 | Johannes<sup>(43)</sup> | 2023 | Evaluation of AI Model for Cephalometric Landmark Classification (TG Dental) | Journal of Medical Systems |
| 23 | Kapoor<sup>(27)</sup> | 2022 | Development, testing, and feasibility of a customized mobile application for obstructive sleep apnea (OSA) risk assessment: A hospital-based pilot study | Journal of Oral Biology and Craniofacial Research |
| 24 | Kim<sup>(44)</sup> | 2019 | DeNTNet: Deep Neural Transfer Network for the detection of periodontal bone loss using panoramic dental radiographs | Scientific Reports |
| 25 | Liu<sup>(45)</sup> | 2020 | A Smart Dental Health-IoT Platform Based on Intelligent Hardware, Deep Learning, and Mobile Terminal | IEEE Journal of Biomedical and Health Informatics |
| 26 | Livas<sup>(46)</sup> | 2019 | Concurrent validity and reliability of cephalometric analysis using smartphone apps and computer software | Angle Orthodontist |
| 27 | Lo Giudice<sup>(47)</sup> | 2021 | A new software architecture proposal for an evidence-based decision support system in dentistry | Minerva Dental and Oral Science |
| 28 | Mazzoli<sup>(48)</sup> | 2017 | Use of MIMICS® software in three-dimensional cephalometric evaluation of soft tissues of the face | Computer Methods in Biomechanics and Biomedical Engineering: Imaging and Visualization |
| 29 | Muhammed Sunnetci<sup>(28)</sup> | 2022 | Periodontal bone loss detection based on hybrid deep learning and machine learning models with a user-friendly application | Biomedical Signal Processing and Control |
| 30 | Najmuddin<sup>(49)</sup> | 2018 | Logicon: A third eye for caries detection | Journal of Indian Academy of Oral Medicine and Radiology |
| 31 | Orhan<sup>(50)</sup> | 2023 | Determining the reliability of diagnosis and treatment using artificial intelligence software with panoramic radiographs | Imaging Science in Dentistry |
| 32 | O'Toole<sup>(23)</sup> | 2019 | Investigation into the validity of WearCompare, a purpose-built software to quantify erosive tooth wear progression | Dental Materials |
| 33 | O'Toole<sup>(22)</sup> | 2020 | Influence of scanner precision and analysis software in quantifying three-dimensional intraoral changes: Two-factor factorial experimental design | Journal of Medical Internet Research |
| 34 | Preus<sup>(51)</sup> | 2015 | A new digital tool for radiographic bone level measurements in longitudinal studies | BMC Oral Health |
| 35 | Reyes Salgado<sup>(29)</sup> | 2022 | Design of Open Code Software to Downs and Steiner Lateral Cephalometric Analysis with Tracing Landmarks | Digital |
| 36 | Roehl<sup>(30)</sup> | 2022 | Tooth Wear Evaluation System (TWES) 2.0—Reliability of diagnosis with and without computer-assisted evaluation | Journal of Oral Rehabilitation |
| 37 | Santeiro-Hermida<sup>(52)</sup> | 2023 | Validation Analysis of Panoramic Dental Application (PDApp) Software as a Tool for Predicting Third Molar Eruption Based on Panoramic Radiograph Images | Applied Sciences (Switzerland) |
| 38 | Sayar<sup>(53)</sup> | 2017 | Manual tracing versus smartphone application (app) tracing: a comparative study | Acta Odontologica Scandinavica |
| 39 | Zadrozny<sup>(21)</sup> | 2022 | Artificial Intelligence Application in Assessment of Panoramic Radiographs | Diagnostics |

n=39
Source: Developed by authors

Table 3 shows the relationship of computer tools described by the articles with the country where they were developed, year of creation, programming language, platforms on which they can be used, the specialty with which they are related, whether they were validated, open source, free, and the payment model to which they are subject. “CephNinja, ”^(46, 53)^ “Diagnocat, ”^(21, 50)^ “OneCeph”^(24, 46)^ y “WearCompare”^(22, 23)^ were the recurrent tools and that 2 (5.1%) articles each described their use. Only six ^(18, 26, 31, 36, 41, 48)^ (15.4%) validation processes were not referred to in the articles, and the rest were validated.

**Table 3.** List of informatic tools extracted from articles with general data on development and availability.

| # | Tool name | Country | Year | Programming language | Platforms | Specialty | Validated | Open code | Free | Payment model |
| --- | --- | --- | --- | --- | --- | --- | --- | --- | --- | --- |
| 1 | 3D Slicer<sup>(34)</sup> | USA | 1999 | C++ | Windows, macOS y Linux. | O | Yes | Yes | Yes | FM |
| 2 | Apox<sup>(17)</sup> | Netherlands | 2022 | NR | NR | RD | Yes | NR | NR | NR |
| 3 | Artificial_Intelligence_Toolbox<sup>(28)</sup> | Turkey | NR | MATLAB 2020b - GUI | Windows, Android | P | Yes | NR | NR | NR |
| 4 | Boneprox<sup>(26)</sup> | Sweden | 2021 | NR | Windows | RD | NR | NR | NR | NR |
| 5 | Cephalopoint<sup>(29)</sup> | Mexico | 2022 | Octave | Android, iOS, Linux<br>Windows, Unix | O | Yes | Yes | Yes | FM |
| 6 | CephNinja<sup>(46, 53)</sup> | USA - India | NR | NR | Android, iOS | O | Yes | NR | NR | NR |
| 7 | CranioCatch<sup>(19)</sup> | Turkey | 2022 | Python (PyTorch) | Android, iOS, Linux<br>Windows, Unix | RD | Yes | No | No | FPM |
| 8 | Decision Support System<sup>(47)</sup> | Italy | NR | Visual C# | NR | RD | Yes | NR | NR | NR |
| 9 | DCDNet<sup>(20)</sup> | Turkey | 2022 | Python (TensorFlow) | NR | RD | Yes | NR | NR | NR |
| 10 | Denti.AI<sup>(25)</sup> | Canada | 2017 | Faster R-CNN, VGG-16 | Android, iOS, Linux<br>Windows, Unix | RD | Yes | No | No | FPM |
| 11 | DeNTNet<sup>(44)</sup> | South Korea | NR | NR | NR | P | Yes | NR | NR | NR |
| 12 | Diagnocat<sup>(21, 50)</sup> | USA | 2017 | Python | Any Web Navigator | OS | Yes | No | No | FPM |
| 13 | DoctOral<sup>(37)</sup> | Italy | 2014 | NR | Android, iOS | OS | Yes | No | Yes | FM |
| 14 | DSDapp<sup>(38)</sup> | Brazil | 2018 | NR | iOS | RD | Yes | No | No | FPM |
| 15 | Endo 10<sup>(32)</sup> | Brazil | 2021 | NR | Android, iOS | E | Yes | NR | Yes | FM |
| 16 | Faster R-CNN<sup>(15)</sup> | Saudi Arabia | 2020 | Python (TensorFlow) | Linux, Windows, macOS | P | Yes | NR | NR | NR |
| 17 | Geomagic<sup>(31)</sup> | USA | 1996 | NR | Windows | OR | NR | No | No | TM |
| 18 | iHome DentalHealth-IoT<sup>(45)</sup> | China | 2018 | NR | Android, iOS | RD | Yes | No | NR | NR |
| 19 | ImageJ (Plugin)<sup>(51)</sup> | Norway | 2015 | NR | Windows, macOS, Linux | P | Yes | Yes | Yes | F |
| 20 | LinguApp<sup>(18)</sup> | Italy | 2020 | JavaScript | Android, iOS | OS | NR | No | Yes | FM |
| 21 | Logicon<sup>(49)</sup> | USA | 2002 | NR | NR | RD | Yes | No | No | NR |
| 22 | MSC<sup>(33)</sup> | Saudi Arabia | 2021 | Python - Yolov5-x | Windows, Linux, macOS. | RD | Yes | NR | NR | NR |
| 23 | Mask-RCNN<sup>(40)</sup> | NR | NR | Python (Tensorflow, Keras) | NR | E | Yes | NR | NR | NR |
| 24 | MeMoSA<sup>(41)</sup> | Malaysia | 2017 | React Native | Android | OS | NR | NR | NR | NR |
| 25 | MIMICS<sup>(48)</sup> | Belgium | NR | C++ | Windows, Linux | O | NR | No | No | FPM |
| 26 | NutriOdonto<sup>(16)</sup> | Brazil | 2018 | NR | Android | RD | Yes | Yes | Yes | FM |
| 27 | OneCeph<sup>(24, 46)</sup> | India | NR | NR | Android | O | Yes | No | Yes | NR |
| 28 | OrthoAnalyzer<sup>(35)</sup> | Denmark | 2014 | NR | NR | O | Yes | No | No | FPM |
| 29 | OSA-Risk Assessment Tool<sup>(27)</sup> | India | 2020 | HTML, PHP | Any Web Navigator | O | Yes | Yes | Yes | FM |
| 30 | PDApp<sup>(52)</sup> | Spain | 2022 | C++ | Linux, Windows | OS | Yes | No | NR | NR |
| 31 | RDFNet<sup>(42)</sup> | China | 2021 | Python (Pytorch) | Windows, Linux, MacOS | RD | Yes | NR | NR | NR |
| 32 | Remote-I<sup>(39)</sup> | Australia | 2015 | NR | Android | RD | Yes | NR | NR | NR |
| 33 | TG Dental<sup>(43)</sup> | NR | NR | NR | NR | O | Yes | NR | NR | NR |
| 34 | TWES<sup>(30)</sup> | Germany | 2020 | MariaDB | Windows, macOS | OR | Yes | No | No | FPM |
| 35 | TSegNet<sup>(36)</sup> | Hong Kong | 2019 | NR | iOS | O | NR | No | No | FPM |
| 36 | WearCompare<sup>(22, 23)</sup> | United Kingdom | 2019 | C++ | Windows, Linux | RD | Yes | Yes | Yes | FM |

n=36
**Source:** Developed by authors - **Caption:** O (Orthodontics), OS (Oral surgery), OR (Oral Rehabilitation), RD (Restorative dentistry), P (Periodontics), E (Endodontics), NR (Not referred), FPM (Full Paid Model), FM (Free Model), F (Freemium), TM (Test Model)

The United States was the most representative country for the development of five (14.3%) of these computer tools (3D Slicer, CephNinja, Diagnocat, Geomagic, and Logicon). (Table 3)

More informatic tools related to "Restorative Dentistry" have been developed than the rest of the specialties of the dental sciences with 14 (40%), followed by the tools related to "Orthodontics" 9 (25.7%) and those useful for "Oral Surgery" 5 (13.9%). (Tables 3 and 4)

**Table 4.** Distribución de herramientas informáticas según especialidad en la que ayudan al diagnóstico.

| RESTORATIVE DENTISTRY | ORTHODONTICS | ORAL SURGERY | PERIODONTICS | PROSTHODONTICS | ENDODONTICS |
| --- | --- | --- | --- | --- | --- |
| Apox | 3D Slicer | Diagnocat | Artifical_Intelligence_Toolbox | Geomagic | Endo 10 |
| Boneprox | Cephalopoint | DoctOral | DeNTNet | TWES | Mask-RCNN |
| CranioCatch | CephNinja | LinguAPP | Faster R-CNN |  |  |
| Decision Support System | MIMICS | MeMoSA | ImageJ (Plugin) |  |  |
| DCDNet | OneCeph | PDApp |  |  |  |
| Denti.Ai | OrthoAnalyzer |  |  |  |  |
| DSDapp | OSA-Risk Assessment Tool |  |  |  |  |
| iHome DentalHealth-IoT | TG Dental |  |  |  |  |
| Logicon | TSegNet |  |  |  |  |
| MSC |  |  |  |  |  |
| NutriOdonto |  |  |  |  |  |
| RDFNet |  |  |  |  |  |
| Remote-I |  |  |  |  |  |
| WearCompare |  |  |  |  |  |

n=36

The most predominant programming language for tool development was Python, with 7 (19.4%), followed by C++, which was used for the development of 5 (13.8%). (Table 3)

Thirty (83.3%) tools were validated, and six (16.7%) were not specified due to lack of information in the articles. A total of six (16.7%) open-code tools were described, another 16 (44.4%) were not, and of the rest, no records of this data were found in the articles. In addition, 10 (27.8%) tools were declared free, 10 (27.8%) were not, and the rest were not provided in the articles. (Table 3)

Table 4 shows the grouping of the tools according to the specialties in which they are useful for diagnosis to facilitate consultation according to the affinity of interested readers.

## DISCUSSION

The search engines used to carry out this research were Scopus, Web of Science, and PubMed, which are essential databases for systematic reviews owing to their wide coverage and unique characteristics. Scopus, as highlighted by Schwager and Schalk^(54)^, demonstrated high sensitivity in retrieving relevant articles, while Web of Science, as shown in Kumpulainen and Seppänen^(55)^, efficiently identified additional studies by searching for citations. PubMed, which is renowned for its extensive coverage of the biomedical literature, is crucial for health-related systematic reviews. The use of these databases ensures a thorough search process, as highlighted in the work of Alfandaria and Taylor^(56)^, who found that replicability varied from platform to platform. Each database contributes to minimizing bias and maximizing the retrieval of relevant studies, aligning with best practices for systematic literature searches.^(57)^ Based on these approaches, the reason why these databases were chosen for the search of articles in this research is shown. The tools described in the results have great potential to improve diagnosis, treatment planning, and information management in the field of dentistry and can be grouped to provide a complete overview and analyze them in detail according to their specialization and functions.

Among the tools for cephalometric analysis and diagnosis, Cephalopoint^(29)^ and OneCeph^(24, 46)^ offer digital cephalometric analysis, facilitating the diagnosis and planning of orthodontic treatments. CephNinja^(46, 53)^ allows digital cephalometric plotting on mobile devices, further streamlining the process. In addition, MIMICS^(48)^ provides three-dimensional cephalometric evaluation of facial soft tissues, which is crucial for assessing facial harmony and orthodontic planning.

Studies by da Fonseca Reis et al.^(58)^ and Prince et al.^(59)^ argue that computer tools that enable cephalometric tracking offer several advantages in orthodontic diagnosis and treatment planning. These tools improve the accuracy, reproducibility, and efficiency of cephalometric image analyses. On the other hand, they eliminate human errors associated with manual tracking, allow automatic detection of landmarks, and offer a high level of agreement with manual methods.^(60)^ In addition, according to Reyes Salgado^(29)^ computerized cephalometric tracing methods are more reliable and consistentthano manual tracing, ensuring accurate measurements for treatment proposals and diagnostic hypotheses.

For the diagnosis of cavities and other dental diseases, we found Logicon,^(49)^ RDFNet, ^(42)^ and DCDNet^(20)^, which specialize in detecting cavities in dental imaging, while Mask-RCNN^(40)^ focuses on the detection and classification of periapical lesions. In addition, Denti.Ai^(25)^ y Diagnocat^(21, 50)^ uses AI to identify a variety of dental pathologies, with Denti.Ai^(25)^ also focusing on implants and crowns. On the other hand, TG Dental^(43)^ classifies malocclusions without manual identification of landmarks using AI to speed up diagnosis and planning.

Computer tools for diagnosing tooth decay from images offer objective verification, aid in doctor-patient communication, teledentistry, and potentially improve diagnostic accuracy and efficiency in the detection of oral diseases.^(61)^ Studies such as Tareq et al. ^(62)^ argue that these applications make it possible to predict dental cavitations from non-standardized photographs with reasonable clinical accuracy, improving access to oralhealthcaree in resource-limited areas. In addition, the use of deep learning in panoramic images makes it possible to accurately detect various tooth-related diseases in real-time, helping to plan treatment in time and reducing the risk of misdiagnosis.^(63)^

To support clinical decisions, we have a Decision Support System"^(47)^ that standardizes the decision-making process in dentistry and offers evidence-based therapeutic options. For specific treatments and risk assessment, Endo 10^(32)^ focuses on diagnosis in endodontics, whereas the OSA-Risk Assessment Tool^(27)^ assesses the risk of obstructive sleep apnea.

The useful tools for teledentistry and remote diagnosis was Remote-I, which facilitates remote screening for oral diseases, ideal for contexts where patients cannot physically visit clinics.^(39)^ Telediagnostic services have demonstrated high accuracy rates, comparable to face-to-face examinations, making them a reliable alternative for clinical support, especially in remote areas.^(64)^

For the evaluation of prosthodontics and prosthesis adjustments, Geomagic^(31)^ is available, a tool that ensures that prostheses such as dentures fit correctly to the models of the patient’s mouth. For research data development and management NutriOdonto^(16)^ it facilitates the management of epidemiological data in oral health, which is crucial for research and diagnosis using the epidemiological method.

In addition, 3D Slicer^(34)^ and WearCompare^(22, 23)^ used 3D models to evaluate tooth movements and adjustments during treatment.

The specialized tools for diagnosis and prevention found in the present study were Apox^(17)^, which analyzes panoramic X-rays providing essential clinical details such as the presence of implants and crowns, the Artificial IntelligenceToolbox^(28)^, which detects periodontal bone loss using AI models, and Boneprox^(26)^, which assesses the risk of osteoporosis through dental X-rays.

Kolokythas et al.^(65)^ argued that AI in dentistry simplifies complex protocols, helps deliver high-quality care, and improves decision-making skills, thus benefiting both doctors and patients. By using large data sets and learning patterns, machine learning can predict disease risks and aid early intervention, which could have a significant impact on patients’ lives.^(66)^ In addition, artificial intelligence applications can help doctors diagnose oral diseases, optimize treatment plans, and improve treatment outcomes, especially in pediatric dentistry.^(67)^

The large number of computer tools found in this study focused on the diagnosis and management of specific diseases. DoctOral^(38)^ focused on the diagnosis of oral lesions. LinguAPP^(18)^ it helps in the diagnosis of soft tissue injuries in the oral cavity. MSC^(33)^ can be used to detect pathological pulp exposure on periapical radiographs. MeMoSA^(41)^ it facilitates the documentation of oral injuries and communication between dentists and specialists. The iHome DentalHealth-IoT^(45)^ detects a wide range of dental problems, from cavities to periodontal diseases. ImageJ (Plugin) ^(51)^ It reduces bias in the measurement of bone loss in radiographic images. PDApp^(52)^ predicts the potential for eruption or retention of third molars using a user-friendly interface for radiological image manipulation. Faster R-CNN^(15)^ Detects gingivitis in orthodontic patients using intraoral imaging. The DSDapp^(38)^ facilitates the planning of multidisciplinary aesthetic treatments. TSegNet^(36)^ Uses Deep Learning for Dental Segmentation in 3D models.

The limitations of the evidence included in the review may be derived from the insufficient information needed regarding each of the tools, such as the validation of some tools. In addition, excessive concentration in certain specialties, such as restorative dentistry, shows a possible limitation in the diversity of areas covered in the research.

## CONCLUSIONS

The incorporation of informatics tools into dental practice has notable benefits in terms of diagnostic accuracy and efficiency in treatment planning. However, there is evidence for the need for a more robust regulatory framework to ensure the proper validation of these technologies before their clinical implementation. In addition, it is crucial to encourage the continuous training of dental professionals in the use of these technologies to maximize their benefits and ensure optimal patient care. This review underlines the importance of further research to explore the full potential of informatics applications in the dental field, particularly with regard to their integration into existing health systems and accessibility in resource-limited areas.

## Data Availability

The database of the articles included in the study with all the details collected according to the variables is located in the Mendeley Data repository (https://doi.org/10.17632/yx9jts8mv7.1), advocating for the reproducibility, sharing and systematicity of data according to the principles of Open Science.

